# Time is of the essence: impact of delays on effectiveness of contact tracing for COVID-19, a modelling study

**DOI:** 10.1101/2020.05.09.20096289

**Authors:** Mirjam E. Kretzschmar, Ganna Rozhnova, Martin Bootsma, Michiel van Boven, Janneke van de Wijgert, Marc Bonten

## Abstract

**Background:** With confirmed cases of COVID-19 declining in many countries, lockdown measures are gradually being lifted. However, even if most social distancing measures are continued, other public health measures will be needed to control the epidemic. Contact tracing via conventional methods or mobile app technology is central to control strategies during deescalation of social distancing. We aimed to identify key factors for a contact tracing strategy (CTS) to be successful.

**Methods:** We evaluated the impact of timeliness and completeness in various steps of a CTS using a stochastic mathematical model with explicit time delays between time of infection and symptom onset, and between symptom onset, diagnosis by testing, and isolation (testing delay). The model also includes tracing of close contacts (e.g. household members) and casual contacts, followed by testing regardless of symptoms and isolation if positive, with different delays (tracing delay) and coverages (tracing coverage). We computed effective reproduction numbers of a CTS (R_cts_) for a population with social distancing measures and various scenarios for isolation of index cases and tracing and quarantine of its contacts.

**Findings:** For the best-case scenario (testing and tracing delays of 0 days and tracing coverage of 80%), and assuming that around 40% of transmission occur before symptom onset, the model predicts that the effective reproduction number of 1.2 (with social distancing only) will be reduced to 0.8 by adding contact tracing. A testing delay of 2 days requires tracing delay to be at most 1 day, or tracing coverage to be at least 80% to keep R_cts_ below 1. With a testing/isolation delay of 3 days, even the most efficient CTS cannot reach R_cts_ values below 1. The effect of minimizing tracing delay (e.g., with app-based technology) declines with decreasing coverage of app use, but app-based tracing alone remains more effective than conventional tracing alone even with 20% coverage. The proportion of transmissions per index case that can be prevented depends on testing and tracing delays, and ranges from up to 80% in the best-case scenario (testing and tracing delays of 0 days) to 42% with a 3-day testing delay and 18% with a 5-day testing delay.

**Interpretation:** In our model, minimizing testing delay had the largest impact on reducing onward transmissions. Optimizing testing and tracing coverage and minimizing tracing delays, for instance with app-based technology, further enhanced CTS effectiveness, with a potential to prevent up to 80% of all transmissions. Access to testing should therefore be optimized, and mobile app technology may reduce delays in the CTS process and optimize contact tracing coverage.

**Research in context:** *Evidence before this study:* We searched PubMed, bioRxiv, and medRxiv for articles published in English from January 1, 2020, to June 20, 2020, with the following keywords: (“2019-nCoV” OR “novel coronavirus” OR “COVID-19” OR “SARS-CoV-2”) AND “contact tracing” AND “model*”. Population-level modelling studies of severe acute respiratory syndrome coronavirus 2 (SARS-CoV-2) have suggested that isolation and tracing alone might not be sufficient to control outbreaks and additional measures might be required. However, few studies have focused on the effects of lifting individual measures once the first wave of the epidemic has been controlled. Lifting measures must be accompanied by effective contact tracing strategies (CTS) in order to keep the effective reproduction number below 1. A detailed analysis, with special emphasis on the effects of time delays in testing of index patients and tracing of contacts, has not been done.

*Added value of this study:* We performed a systematic analysis of the various steps required in the process of testing and diagnosing an index case as well as tracing and isolating possible secondary cases of the index case. We then used a stochastic transmission model which makes a distinction between close contacts (e.g. household members) and casual contacts to assess which steps and (possible) delays are crucial in determining the effectiveness of CTS. We evaluated how delays and the level of contact tracing coverage influence the effective reproduction number, and how fast CTS needs to be to keep the reproduction number below 1. We also analyzed what proportion of onward transmission can be prevented for short delays and high contact tracing coverage. Assuming that around 40% of transmission occurs before symptom onset, we found that keeping the time between symptom onset and testing and isolation of an index case short (<3 days) is imperative for a successful CTS. This implies that the process leading from symptom onset to receiving a positive test should be minimized by providing sufficient and easily accessible testing facilities. In addition, reducing contact-tracing delays also helps to keep the reproduction number below 1.

*Implications of all the available evidence:* Our analyses highlight that CTS will only contribute to containment of COVID-19 if it can be organised in a way that time delays in the process from symptom onset to isolation of the index case and his/her contacts are very short. The process of conventional contact tracing should be reviewed and streamlined, while mobile app technology may offer a tool for gaining speed in the process. Reducing delay in testing subjects for SARS-CoV-2 should be a key objective of CTS.

## Introduction

As the first wave of the SARS-CoV-2 has reached its peak of cases in many countries, societies are preparing so-called exit-strategies from the COVID-19 lockdown, while still successfully controlling transmission. Contact tracing, in combination with quarantine and potentially testing of the contacts, is considered a key component in a phase when lockdown measures are gradually lifted^1-8^. Contact tracing is an intervention, where an index case with confirmed infection is asked to provide information about contact persons, who were at risk of acquiring infection from the index case within a given time period before the positive test result. These contact persons are then traced and informed about their risk, quarantined, and tested if eligible for testing according to national testing guidelines. This requires upscaling of conventional contact tracing capacity. The potential of mobile apps to support contact tracing is widely discussed and such technology has been used in several Asian countries. Although these countries have successfully reduced case numbers, no causal relationship between use of app technology and epidemic control has yet been shown^9-14^. Many uncertainties remain on the optimal process of contact tracing with conventional methods and/or mobile applications, on the timing of testing for current or past infection, and on the required coverage of contact tracing needed.

Modelling studies have demonstrated how mobile applications can increase effectiveness of contact tracing, compared to conventional approaches for contact tracing, but effectiveness depends on what proportion of the population will use the app consistently for a sufficiently long period of time^9^. Modelling studies have predicted that contact tracing alone cannot control an outbreak if tracing coverage is too low^2,15^. What tracing coverage is needed depends on how much transmission occurs before symptom onset, and on the details of the tracing process.

In previous work, we have investigated the impact of timeliness and completeness of case reporting for the effectiveness of surveillance and interventions^16,17^, and we quantified the timeliness of contact tracing of infected passengers during an airline flight for the 2009 pandemic influenza^18^. In all of these studies, the timing of various steps in the monitoring and intervention chain emerged as one of the key factors for effectiveness of a public health response. Usually, there are identifiable delays in the response chain that may be critical to the overall effectiveness of a strategy.

Here we analyze in detail the process chain of identifying index cases by symptom-reporting followed by testing, and subsequent contact tracing, with the aim to inform policy makers on the relative importance of key steps in the process. We use a mathematical model that reflects the various steps and delays in the contact tracing process to quantify the impact of delays on the effective reproduction number and the fraction of onward transmission prevented per diagnosed index case^5,19^.

## Methods

### Time delays in contact tracing

Our starting point is an assumed effective reproduction number (*R*_e_) for COVID-19 of around 1, describing a situation with “social distancing but measures lifted to some extent”. We then quantify the relative contribution of the individual components of a contact tracing strategy (CTS) required to bring and maintain the effective reproduction number with CTS (*R*_CTS_) to a value below 1. For simplicity we do not include transmission in healthcare settings, as in healthcare settings like nursing homes, which can be viewed as closed populations, other interventions are more appropriate.

We break down the process of contact tracing in two steps (Figure 1; Supplementary Information Table S1).

**Figure 1:**
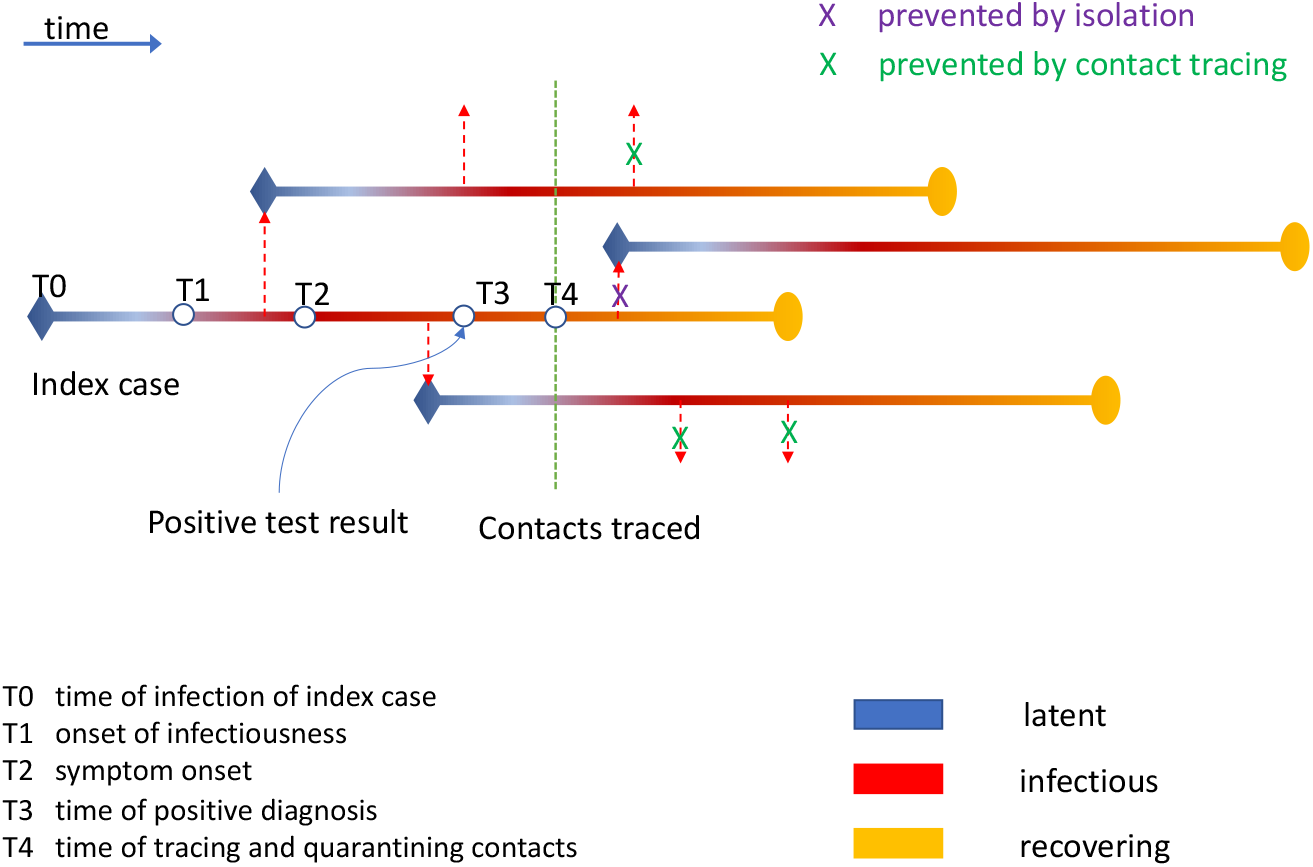
Schematic of the contact tracing process and its time delays.

- An index case acquires infection (at time T_0_), then after a short latent period becomes infectious (at time T_1_), and finally symptomatic (at time T_2_), which is here defined as “being eligible for testing”. Subsequently a proportion of all symptomatic subjects gets tested and diagnosed (at time T_3_). The time between T_2_ and T_3_ is called the “testing delay” (D_1_ = T_3_ - T_2_), and may vary between 0 and 7 days, and in this period individuals might self-quarantine. We refer to the proportion of all symptomatically infected cases that are tested as testing coverage and vary it from 20% to 80% in increments of 20%. After being diagnosed, we assume index cases are isolated with no further transmission.
- The second step is tracing contacts of the index case, which occurs at time T_4_. A fraction of those contacts will be found and tested. We assumed that all contacts, regardless of symptoms, are offered testing, and that those testing negative do not spread. Those who are found infected will be isolated, with effectiveness ranging from 0% to 100%. We assume that contacts in isolation do not spread. The time between T_3_ and T_4_ is the “tracing delay” (D_2_ = T_4_ – T_3_), which may range from 0 (for instance with app technology) to 3 days (with conventional approaches). In this step, tracing coverage is defined as the proportion of contacts detected, which either depends on the capacity of conventional approaches (ranging from 20% to 80% in increments of 20%) or on the fraction of the population using suitable app technology for screening (ranging from 20% to 100% in increments of 20%). We did not consider hybrid approaches of combined conventional and app-based CTS.

The best-case scenario we consider is that persons eligible for testing are immediately tested (coverage 80%) with a very fast test result (test-delay 0 days) and immediate isolation when testing positive, followed by immediate tracing (trace delay 0 days) of all contacts, that immediately adhere to isolation measures (coverage 80%). We consider more realistic scenarios where testing and tracing are suboptimal, e.g. a conventional CTS, and we vary these parameters separately in a sensitivity analysis (see Supplementary Information).

### Impact on effectiveness on population level

To analyse the impact of these time delays on the effectiveness of contact tracing we use a model introduced by Kretzschmar et al^19^, which was adapted for SARS-CoV-2^5^. The stochastic model describes an epidemic as a branching process with progression through latent infection and infectious period in time steps of 1 day. Infectivity and probability of symptom onset per day of the infectious period, and numbers of contacts per day were fitted to distributions taken from published data. ^20-24^ We distinguish between close contacts (e.g. household contacts, but also other high-risk contacts) and casual contacts, which differ in the risk of acquiring infection from the index case. Also, the time required for tracing and isolating infected contacts and the coverage of tracing may differ between these types of contacts and between different CTS (i.e., conventional contact tracing versus mobile app supported contact tracing). We assume that isolation is perfect, i.e. that isolated persons do not transmit any longer, and that all traced infected contacts are isolated, regardless of whether they develop symptoms or not. The model allows for explicit computation of the basic reproduction number R_0_, the effective reproduction number under social-distancing interventions R_e,_ and the effective reproduction number with CTS (R_cts_). Reproduction numbers were calculated as expectations, and distributions of individual reproduction numbers were simulated. The model was coded in Mathematica 12.1. For details, see the Supplementary Information.

### Parameter settings

We assumed that without social distancing individuals have on average 4 close contacts per day and around 9 casual contacts per day, with stochastic variability. The distributions were fitted to data from the Polymod study for the Netherlands^23^. Transmission probability per contact for close contacts was taken to be 4 times higher than for casual contacts. Symptomatic and asymptomatic cases were assumed to be equally infectious. Overall, the transmission probability was calibrated to a basic reproduction number of R_0_ = 2.5. For social distancing, we assumed that close contacts were reduced by 40% and casual contacts by 70%. The resulting effective reproduction number was R_e_ = 1.2.

### Uncertainty of model outcomes

We considered uncertainty due to stochastic variability, and uncertainty due to possible variation in parameter estimates. We dealt with stochastic variability by computing individual reproduction numbers for 1000 individuals for all scenarios, and plotted their distributions as boxplots. Parameter uncertainty was explored by performing simulations using hypercube sampling for transmission probabilities and probabilities of symptom onset per day of the infectious period (Supplementary Information).

### Scenarios modelled

We analyzed the impact of various testing and tracing delays and tracing coverage on the effective reproduction number R_cts_ while keeping the testing coverage at 80%. For comparison, we also considered the strategy where symptomatic individuals get tested and isolated, without subsequent tracing (R_iso_). We varied the testing delay D_1_ between 0 and 7 days, the tracing delay D_2_ between 0 and 3 days, and tracing coverages between 0% and 100% in increments of 20%. For conventional contact tracing, we assumed that coverage is higher for close contacts than for casual contacts.

We then compared the effectiveness of conventional CTS alone with a scenario in which mobile app technology is used for alerting subjects to be tested and for tracing contacts. Differences between these strategies were taken as follows. The testing delay (D_1_) is reduced with app technology. We assumed a conventional CTS setting in which symptomatic individuals need to decide to seek health care to get tested, and we assumed that with app technology symptomatic persons get alerted and can be tested without health care interference. For conventional CTS we assumed suboptimal coverage in identifying contacts from the week before diagnosis due to recall bias, especially for casual contacts. For CTS with mobile app technology we assume 60% and 80% tracing coverage of the contacts of subjects using app technology. We show also results for 100% coverage, although realistically more than 80% is not feasible, because not all contacts may be correctly identified and compliance with isolation of those tested positive may not be perfect. We assume that tracing goes back for 7 days before the positive test result. The exact parameter values for this comparison are shown in Table 1.

**Table 1:** Comparison Conventional CT and Mobile app CT.

|  | **Conventional CT** | **Mobile app CT** |
| --- | --- | --- |
| **Testing coverage** | 80% | 20%, 40%, 60%, 80%, 100% |
| **Testing delay ($D_1$), assuming immediate isolation when testing positive** | 4 days | 0 day |
| **Time to trace close contacts ($D_2$)** | 3 days | 0 day |
| **Time to trace other contacts, assuming testing and isolation of those who test positive** | 3 days | 0 day |
| **Tracing coverage close contacts** | 80% | 20%, 40%, 60%, 80%, 100% |
| **Tracing coverage casual contacts** | 50% | 20%, 40%, 60%, 80%, 100% |
| **Time traced back** | 7 days | 7 days |

Next, we quantified the impact of coverage of testing and app use on the effectiveness of CTS. We varied the percentage of app users in the population between 20% and 100% in increments of 20%. We first considered the situation that testing is provided for 80% of persons with symptoms independent of app use, and app use only influences the fraction of contacts that are traced. Alternatively, assumed that only app users are tested (i.e. testing coverage varies between 20% and 100% in increments of 20%), and coverage of tracing also depends on fraction of app use. In all cases, a contact person could only be traced if both the index case and the contact person were app users, i.e. the probability of a contact being traced is given by the square of the proportion of app users.

Finally, we quantified the fraction of transmissions of an index person that can be prevented, and the contribution to the fraction prevented from isolation and from tracing contacts with decreasing delays. The number of onward transmissions of an index case is by definition described by the effective reproduction number of the realized scenario. Therefore, the difference of reproduction numbers between two intervention scenarios under the condition that an index case is diagnosed, describes the fraction of onward transmissions prevented. For contact persons, this is the fraction of the total infectivity that lies after the time of isolation, i.e. the part of infectiousness that is prevented by contact tracing. In other words, a contact person who is detected and isolated before the start of their infectious period is a fully prevented transmission, while a contact person who is only traced and identified after 70% of their infectivity has passed, is counted as 0.3 of a prevented onward transmission.

## Results

In the best-case scenario, if 80% of infectious persons that develop symptoms are tested and isolated within 1 day after symptom onset the effective reproduction number R_e_ is expected to decline from 1.2 to R_iso_ = 1.0, without contact tracing (Figure 2). Contact tracing may further decrease the reproduction number to R_cts_=0.8 in the best case scenario. In the best case scenario – a testing delay of 0 days, a tracing delay of 0 days, and a tracing coverage of 80%, the additional reduction of R_cts_ predicted by the model is 33%. Yet, with a testing delay of 2 days, tracing delay should be at most 1 day, or tracing coverage should be at least 80% to keep R_cts_ below 1. In these scenarios, the reduction of R_cts_ compared to the best-case scenario is estimated at 17% (Supplementary Information Figure S4). With a testing delay of more than 3 days, even perfect contact tracing cannot bring R_cts_ values below 1.

**Figure 2:**
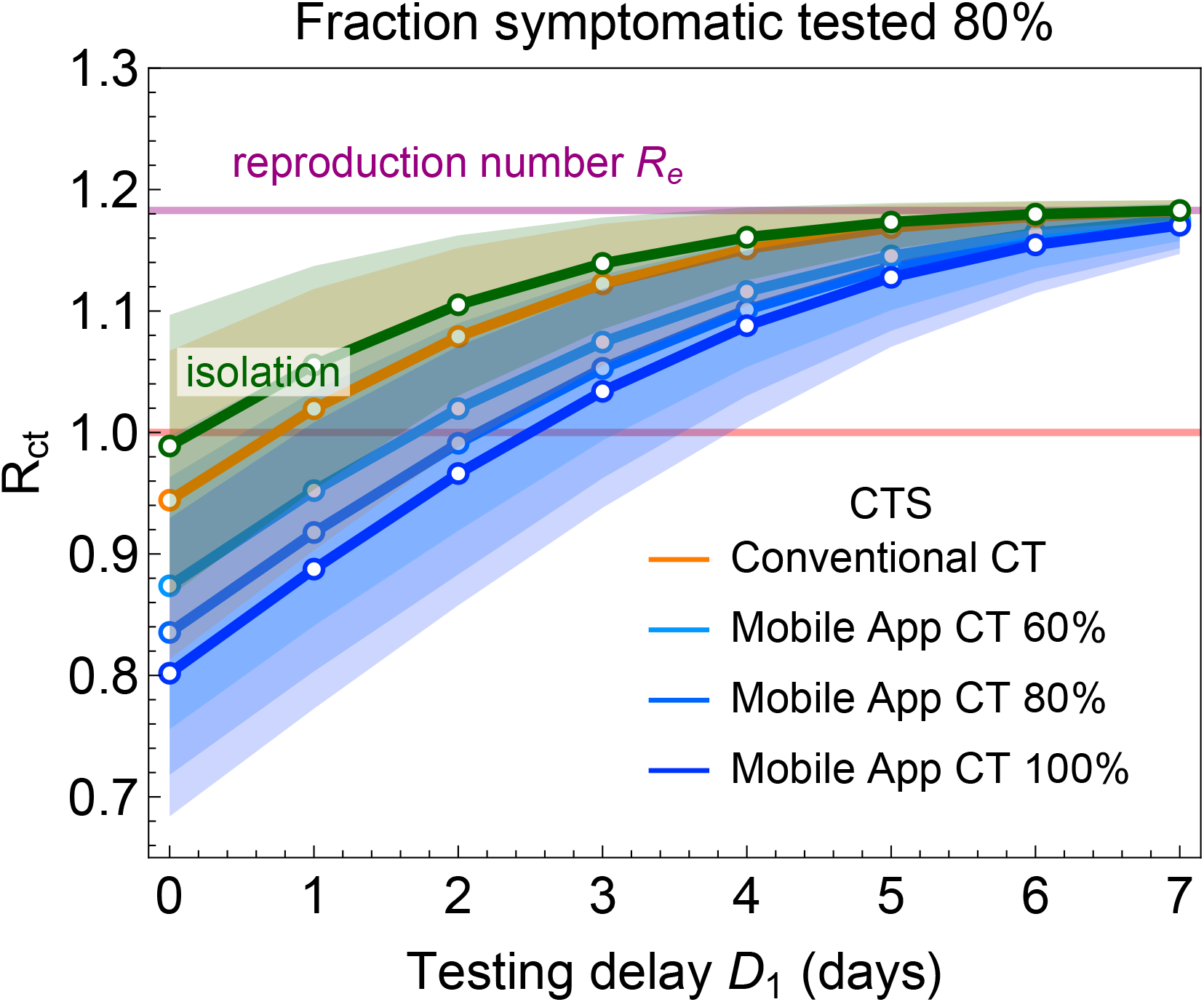
Comparison of a conventional and mobile app CTS. For parameter values, see table 1. We assumed that testing coverage is 80% for the conventional CTS and 60%, 80%, and 100% for the mobile app CTS. For mobile app CTS it is assumed that the tracing coverage equals the testing rate, i.e. it is 60%, 80%, and 100%, respectively. Expected reproduction numbers are shown as a function of testing delay D1.

We assumed that conventional CTS has longer tracing delay and lower tracing coverage than CTS based on app technology which results in marked differences in R_cts_ for the whole range of testing delay (Figure 2A). With conventional CTS, R_cts_ would remain above 1, if the testing delay exceeds 0 days, whereas contact tracing based on app technology could still keep R_cts_ below 1, as long as testing and tracing coverage would be at least 80%, or if testing delay is 1 day and tracing coverage 60%. If the testing delay reaches 5 days or more, app technology adds little effectiveness to conventional CTS or just isolating symptomatic cases.

The reductions of R_e_ (based on social distancing) achieved by isolation of symptomatic cases only, conventional CTS, and mobile app-based CTS are shown in figure 3A. For isolation only and for conventional CTS we assumed a delay of 4 days between symptom onset and isolation of the index case. The relative reductions are independent of the level of R_e_, similar reductions are seen for R_0_, i.e. in a situation without social distancing (Supplementary Information). Conventional CTS, even if applied for all infected subjects with symptoms, is 27% less effective than mobile app-based CTS alone, due to longer tracing delays and lower tracing coverage. Figure 3B shows the distributions of individual reproduction numbers for the testing delays assumed in Table 1, i.e. 4 days for isolation and conventional CT, and 0 days for app-based CTS. Only for app-based CTS the means of the individual reproduction numbers are below 1.

**Figure 3:**
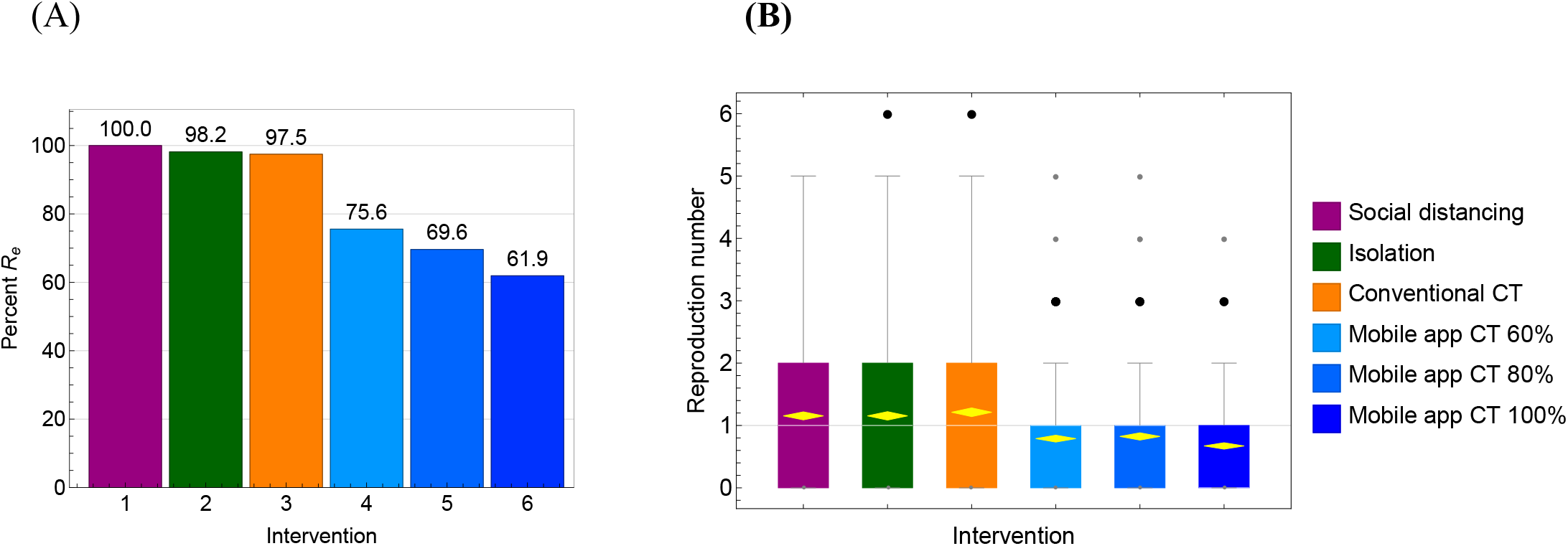
The reduction of the effective reproduction number for various CTS. (A) The reproduction number with CTS, Rcts, is shown as a percentage of the reproduction number where only social distancing is implemented (Re). For the isolation scenario and conventional tracing scenario we assumed that there is a delay of 4 days between symptom onset and isolation of the index case. For the mobile app CTS, testing delay was assumed to be 0 days. Testing coverage was assumed to be 80% in the isolation and conventional CT scenarios; app use prevalence was assumed to be 60%, 80%, and 100% in the mobile app CTS. (B) Distributions of individual reproduction numbers for 1000 individuals and the same scenarios as in (A).

The effectiveness of app-based technology declines with lower fractions of persons using it (Figure 4). Yet, app-based tracing on its own remains more effective than conventional tracing alone, even with 20% coverage, due to its inherent speed. Even with low coverage there is a reduction of R_e_, due to fast tracing of a small part of the population. Depending on R_e_, such an approach might be sufficient to reduce R_cts_ to levels below 1. This can be seen in the distributions of the individual reproduction numbers (Figure 4B and 4D), where in 4B the means of the distributions are below 1 for 40% and more app use, while in 4D this is the case for 60% and above.

**Figure 4:**
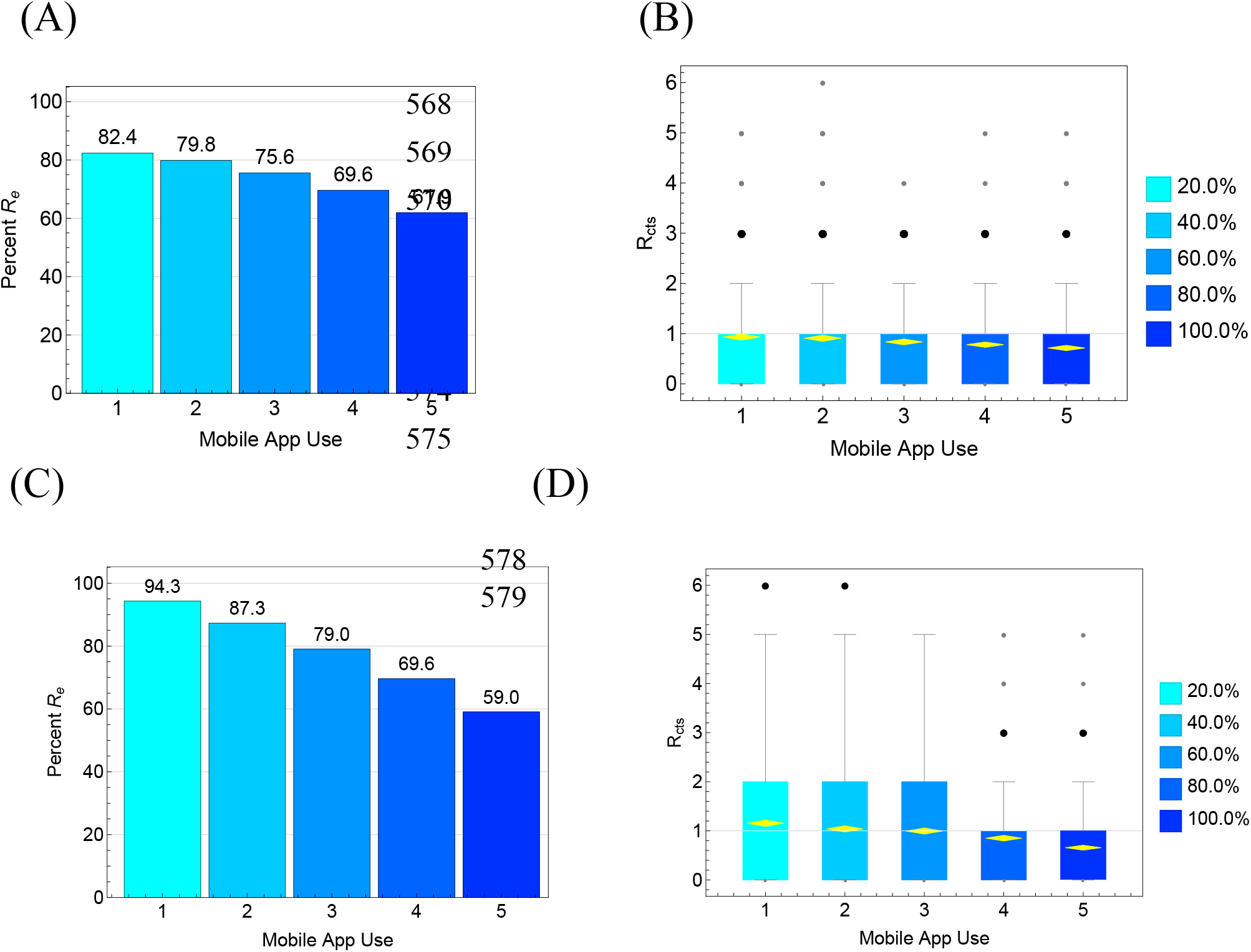
The impact of mobile app use on Rcts for varying levels of app use. In 4A and 4B, we assume that there is also testing of those who do not use the mobile app, so app use only is used for tracing contacts. In 4C and 4D, only app users, who develop symptoms, are tested. Panels A and C show percentage reductions of Re achieved by the CTS; panels B and D show the impact of various CTS on distributions of individuals reproduction numbers.

In Table 2, we quantified proportions of transmissions per index case that can be prevented depending on testing delay, stratified by of isolation of index cases and tracing delays. In the best-case scenario (testing and tracing delay being 0 days) around 80% of transmissions can be prevented if tracing coverage is 80%.

**Table 2.** Percentage of onward transmissions prevented per diagnosed index case for various interventions: only isolation of the index case (left column) or isolation of the index case with tracing and isolation of 80% of infected contacts (columns 2-5).

| **Delay D<sub>1</sub> (days)** | **Isolation only** | **Contact tracing**<br>**Delay D<sub>2</sub> (days)** |  |  |  |
| --- | --- | --- | --- | --- | --- |
|  |  | **3** | **2** | **1** | **0** |
| **0** | 50.4 | 62.4 | 67.8 | 73.9 | 79.9 |
| **1** | 35.7 | 47.3 | 53.4 | 60.7 | 68.5 |
| **2** | 23.4 | 33.0 | 38.9 | 46.5 | 55.4 |
| **3** | 14.2 | 21.0 | 26.0 | 32.9 | 41.8 |
| **4** | 7.8 | 11.9 | 15.7 | 21.4 | 29.1 |
| **5** | 3.8 | 5.9 | 8.4 | 12.5 | 18.4 |
| **6** | 1.6 | 2.4 | 3.8 | 6.4 | 10.4 |

## Discussion

Using a mathematical model that describes the different steps of CTS for COVID-19 we have quantified the relevance of delays and coverage proportions for controlling SARS-CoV-2 transmission. We conclude that reducing the testing delay, i.e. shortening the time between symptom onset and positive test result (assuming immediate isolation), is the most important factor for improving CTS effectiveness. Reducing the tracing delay, i.e. shortening the time of contact tracing (assuming immediate testing and isolation if positive), may further enhance CTS effectiveness. Yet this additional effect rapidly declines with increasing testing delay. The effectiveness of app-based CTS declines with lower app use coverage, but it remains more effective than conventional contact tracing even with lower coverage, due to its inherent speed. CTS therefore has the potential to control virus transmission, and to enable alleviation of other control measures, but only if all delays are maximally reduced. It should be noted that we simulated two CTS systems (conventional CTS with testing and tracing delays and app-based CTS without delays) and ignored hybrid approaches. At present, most European countries are using conventional CTS, but are attempting to reduce delays (for example, by improving testing and tracing capacity and by removing testing barriers), and are piloting or planning the addition of app-based contact-tracing. Such hybrid CTS systems would fall somewhere between the fully conventional and app-based scenarios described in this paper.

Several factors can reduce CTS effectiveness, such as large proportions of cases who remain asymptomatic or are otherwise not diagnosed, and large proportions of contacts who cannot be traced. App-based technology could increase the proportion of tracable contacts, because it does not rely on recall of names and contact details, but this would require the participation of a substantial proportion of the population. App use acceptance may be hampered by privacy concerns and other ethical considerations, which limit its acceptance. Also, app use needs to continue over a long time period, requiring sustained adherence by app users. Low participation does not render CTS useless, however, because it could help to locally extinguish clusters before they grow larger. In addition, every measure that lowers the effective reproduction number, even if it is already below 1, will lower the cumulative case number and speed up the time until elimination of the virus from the population.

The strength of our approach is that it explicitly takes many details of the contact tracing process into account, such that the key factors can be identified. A limitation of our approach is that it does not take population age-structure into account, which may influence the proportion of asymptomatic cases and mobile app use coverage. Also, the willingness of a case to self-isolate depends on age and social norms, may depend on socio-economic status, and is affected by perceived benefit of isolation in relation to perceived risk of the infection to others^25^. We also excluded other heterogeneities while assuming homogeneous mixing^26,27^, and assumed homogeneously distributed use of app technology for different coverage levels. Clustering of non-users may have consequences for overall effectiveness of CTS, similar to clustering of non-vaccinated persons. Furthermore, we ignored that a sizeable portion of transmissions may be acquired nosocomially when population prevalence is still low. ^28^ The model also ignores that some contacts of the index case may have self-quarantined with symptoms before they are traced by CTS, which lowers the benefits of CTS.

Our results add to results from other modelling studies, which showed that CTS can be an effective intervention if tracing coverage is high and if the process is fast^2,15^. A determining factor is the proportion of transmissions occurring before symptom onset, which determines the urgency of tracing and isolating contacts as fast as possible. Our study showed in detail what the role is of each step in the CTS process in making it successful. Our model differs in that it makes a distinction between close and casual contacts, and that we consider scenarios for conventional CTS and mobile app-based CTS characterized by specific delays and coverages.

Our finding of the crucial importance of the first step of CTS, establishing a diagnosis in cases with symptoms, has important consequences. It requires an infrastructure for testing, that allows persons with symptoms to be tested, preferably, within one day of symptom onset. Studies have demonstrated that viral shedding in the respiratory tract is highest at the start of symptoms^29,30^, so early testing will also increase the sensitivity of this approach. To further enhance effectiveness, as many infectious persons as possible need to be tested regardless of symptoms, which requires a low threshold for testing. As the clinical symptoms of COVID-19 are mostly mild and heterogeneous, many persons should be eligible for testing, resulting in a large proportion of negative test results. Future work should determine the optimal balance between the proportion of test-negatives and the effectiveness of CTS.

Our findings also provide strong support to optimize contact tracing. In the Netherlands, CTS was based on establishing contact between an index case and a public health officer, followed by an interview after which contacts are traced. This procedure is labor intensive, time consuming, prone to recall bias, incomplete (anonymous contacts cannot be traced), and usually takes several days. Optimizing this process by improving testing and tracing capacity, removing testing barriers, and by adding app-based and/or other digital technologies to minimize tracing delay are needed to establish optimal control of transmission. These improvements are currently being implemented or considered. Overall, our findings suggest that optimized CTS, with short delays and high coverage for testing and tracing could substantially reduce the reproduction number, which would allow alleviation of more stringent control measures.

## Data availability

The Mathematica code used for the analysis are available on Github under https://github.com/mirjamkretzschmar/ContacttracingModel

## Data Availability

No data were used for this study.

## Contributors

MEK and MB conceived the study. MK designed and programmed the model, and produced output. MvB, MCJB, and GR helped with the analysis and literature research. JvdW contributed to data interpretation and writing. All authors interpreted the results, contributed to writing the manuscript, and approved the final version for submission.

## Acknowledgements

MEK received funding from ZonMw projects number 91216062 and number 10430022010001. GR received funding from Fundação para a Ciência e a Tecnologia project reference 131_596787873. MB received funding from EU H2020 grant RECOVER (H2020-101003589).

We thank Patricia Bruijning-Verhagen and Hans Heesterbeek for useful discussions. This work forms part of RECOVER (*Rapid European COVID-19 Emergency Response research*). RECOVER is funded by the European Union’s Horizon 2020 research and innovation programme under grant agreement No 101003589.

